# Democratizing Infectious Disease Modeling: An AI Assistant for Generating, Simulating, and Analyzing Dynamic Models

**DOI:** 10.1101/2024.07.17.24310520

**Authors:** Joshua L. Proctor, Guillaume Chabot-Couture

## Abstract

Understanding and forecasting infectious disease spread is pivotal for effective public health management. Traditional dynamic disease modeling is an essential tool for characterization and prediction, but often requires extensive expertise and specialized software, which may not be readily available in low-resource environments. To address these challenges, we introduce an AI-powered modeling assistant that utilizes advanced capabilities from OpenAI’s latest models and functionality. This tool enhances the accessibility and usability of infectious disease models and simulation frameworks by allowing users to generate or modify model configurations through intuitive natural language inputs or by importing explicit model descriptions. Our prototype integrates with an established open-source disease simulation framework called the Compartmental Modeling Software (CMS) to provide a seamless modeling experience from setup to analysis. The AI assistant efficiently interprets disease model parameters, constructs accurate model files, executes simulations in a controlled environment, and assists in result interpretation using advanced analytics tools. It encapsulates expert knowledge and adheres to best practices to support users ranging from novices to expert modelers. Furthermore, we discuss the limitations of this AI assistant, particularly its performance in complex scenarios where it might generate inaccurate specifications. By enhancing the ease of disease modeling and supporting ongoing capacity-building initiatives, we believe that AI assistants like this one could significantly contribute to global health efforts by empowering researchers, especially in regions with limited resources, to develop and refine their disease models independently. This innovative approach has the potential to democratize disease modeling in global health, offering a scalable solution that adapts to diverse needs across a wide-range of geographies, languages, and populations.

## 1 Introduction

Infectious diseases continue to pose a significant global health challenge, contributing substantially to morbidity and mortality worldwide. This burden disproportionately affects low- and middle-income countries (1; 2). In these regions, the capacity to implement effective public health interventions—which can vary widely depending on the pathogen—is often constrained by limited resources (3). Despite the availability of effective interventions, the scarcity of resources in high-burden areas complicates efforts to manage and mitigate the impact of diseases effectively (2). Central to overcoming these challenges is planning and evidence-based decision-making, which heavily relies on data collection, quantitative modeling, and analysis (4). However, the detailed modeling and analysis requires specialized skills, knowledge of advanced software, and significant time investment to tailor models to specific geographic contexts (5). This paper explores how recent advancements in artificial intelligence (AI) can address these obstacles, providing specialized tools that empower public health officials to directly engage with modeling and analysis using their own data and hypotheses. By enhancing the accessibility and efficiency of these processes, AI innovations offer a promising avenue for strengthening global health responses in resource-limited settings.

A wide array of mathematical and statistical models are employed in global health for policy planning and disease management (6). Advances in data-driven forecasting methods have enhanced our ability to characterize epidemiological trends and predict future infection trajectories (7). Throughout the COVID-19 pandemic, an international consortium of modelers utilized diverse methodologies to integrate new data streams, refine models, and generate predictions that informed critical public health decisions (8; 9; 7) with varying levels of predictive success (10; 11). Similarly, mechanistic models, which incorporate geography-specific factors such as demographics, human mobility, and historical disease data, have proven invaluable in understanding localized disease scenarios and planning targeted interventions for malaria control and elimination (12; 13). Both types of models—data-driven and mechanistic—demand substantial expertise and development effort. Although a vast number of models and simulation engines have been detailed in scholarly articles and books, with code often shared on platforms like GitHub, utilizing these resources to develop new models or adapt existing ones remains a formidable challenge. Even setting up the computational environment with the correct libraries and packages can be prohibitive. Reducing these barriers to entry is crucial, especially for individuals in resource-constrained environments who may lack the resources or time to develop models from scratch. Facilitating easier access to and use of these modeling tools could significantly accelerate disease modeling efforts in areas most in need.

Recent advances in generative artificial intelligence have catalyzed transformative changes across various scientific and industrial sectors (14; 15). The introduction of OpenAI’s GPT-4 has sparked a myriad of innovative applications leveraging these sophisticated large language models (LLMs) (16). These LLMs excel not only in engaging dialogues but also in a range of functions, including software development, text summarization, advanced data analytics, and the execution of specialized functions—all through natural language prompts—and potentially even general reasoning (16; 15; 17). Critically, these capabilities can be enhanced by the integration of retrieval-augmented generation techniques, which dynamically incorporate external, contextually relevant information into the generation process, thereby enriching the output’s quality and applicability (18). The potential of these LLMs to streamline, enhance, and accelerate complex workflows is substantial, indicating their pivotal role in advancing current technological capabilities.

In this article, we explore the application of these transformative AI tools to develop an AI assistant designed to facilitate infectious disease modeling. We detail the process by which the AI modeling assistant can interpret disease model descriptions—either through direct user prompts or by processing input documents—to generate syntactically correct model files. Subsequently, we illustrate how these models can be simulated using OpenAI’s custom functions and a known simulation engine, and analyze the resultant data using OpenAI’s advanced Code Interpreter capabilities. Furthermore, we identify and discuss the principal limitations of the current prototype and propose potential directions for future development. This discussion aims to provide a comprehensive understanding of how AI can enhance disease modeling practices, particularly for policymakers in settings constrained by resources and expertise.

## 2 Methods

In this section, we describe the methods, models, data, and workflow. Figure 1 portrays both the types of questions that users may ask the assistant and how the AI modeling assistant engages with different elements of the modeling workflow. Note that the AI nodeling assistant does not simply execute the entire modeling workflow, but can be used for only one part of the workflow or iterate between elements adaptively and as the user needs.

**Figure 1:**
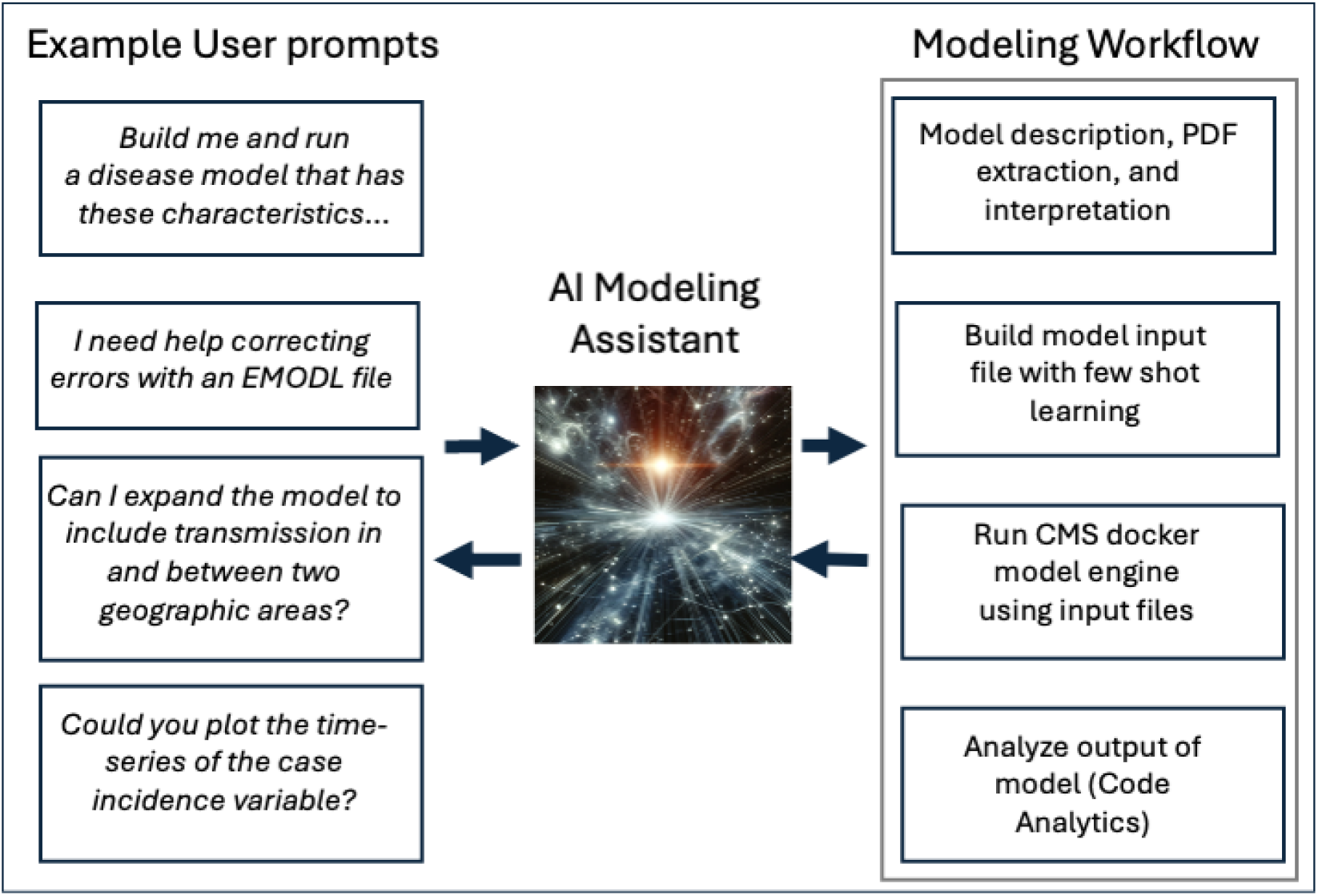
Schematic representation of the AI Modeling Assistant workflow. This diagram illustrates the interactive process between the user and the AI assistant. User prompts (left) initiate various tasks such as building, modifying, or analyzing disease models, which are handled by the AI Modeling Assistant. The assistant’s role involves extracting and interpreting model descriptions from documents, building and running models through a containerized CMS engine, and analyzing the model outputs using advanced Code Interpreter tools. Each step in the workflow demonstrates the assistant’s capabilities in facilitating an iterative, inefficient, and user-friendly approach to disease modeling. The image representing the AI Modeling Assistant was created using OpenAI’s multimodal model.

### 2.1 LLM and computational environment

OpenAI’s Python package, `openai=1.9.0`, was employed to access the API to query the GPT-4 model, specifically version `gpt-4-1106-preview`. The model was utilized with its default parame-ters, including the temperature setting. The functionality of the AI assistant was integrated within this framework. For execution and documentation of the code, a Jupyter Notebook environment was utilized. This environment was managed using Anaconda (conda 23.7.4). The setup included capturing all installed packages and their dependencies. Detailed information regarding the configuration of the computational environment and a complete list of the software and libraries can be found at (19). The jupyter notebook was run on a Apple MacBook Pro with a Apple M3 Max with 36 GB of memory on the Sonoma 14.4.1 macOS.

### 2.2 Prompt Engineering for the AI modeling assistant

The AI modeling assistant is provided a specific role definition. This is directly input into the OpenAI API role definition for the assistant. The prompt describes the AI modeling assistant as responibile to help build the model syntax file, write the file, call the modeling simulation engine, and use Code Interpreter to plot the output data of the CMS; see Appendix §A for the specific role definition. To test the performance of the assistant with few-shot learning, three model files are included in the prompt as examples for the assistant to better understand the context; these model files can be found in Appendix §B. For the AI modeling assistant, we enable the File Retrieval and Code Interpreter functionality offered in the assistant API allowing documents to be uploaded and output data to be analyzed and visualized.

### 2.3 Compartmental Modeling Software for disease simulation

The Compartmental Modeling Software (CMS) is a professionally developed and tested code-base to simulate infectious disease models in the eradication regime using discrete stochastic reaction numerical methods (20). In this disease transmission regime, numerical methods are required that can handle small discrete case counts. The application requires a model description in the format of text and a custom model file format called EMODL. In addition, a configuration file in a json format includes details about the simulation including numerical solver, duration of the simulation, number of realizations, etc. The code respository for CMS can be found at (21) along with links to a docker image containing the application (22). In the workflow described here, we leverage the Docker image and Docker platform for Mac to run the container. The output file from the CMS simulation is automatically uploaded to the thread through the file retrieval functionality.

### 2.4 Code Interpreter for interrogating model output

The Code Interpreter function is enabled for this assistant. The output of the CMS simulation is uploaded in the thread. Code Interpreter is used to analyze and plot the output data from the simulation using natural language prompts.

### 2.5 Defining custom functions

Custom functions are defined to support the AI modeling assistant’s goals. Multiple custom functions are constructed to write the model file from the prompt and to run the CMS simulation using the model file and config file. With the CMS container running in the background, the custom function executes a docker command function that points to the model and configuration file.

## 3 Results

### 3.1 Zero-shot learning and prompt engineering can produce inaccurate model files

Using the assistant’s role description alone, it is possible to prompt the AI to generate an EMODL file based on a specified disease description. However, the results indicate that the syntax of the EMODL files generated from nearly every model query was incorrect, as demonstrated in the example provided in Appendix §C. While the pre-trained model seems to possess a foundational understanding of the CMS framework and EMODL format, the files it produced were often flawed. For the example in Appendix §C, the EMODL file is incomplete (does not include the import packages and the start-model and end-model flags), inaccurate (does not define the species, params, or reaction correctly), and inconsistent (does not replicate the reaction definition correctly).

### 3.2 Few-shot learning and prompt engineering produces model files from natural language prompts

Incorporating three EMODL files from a public GitHub repository into the prompts as examples significantly improved the accuracy of the AI-generated model files. Notably, the syntax of the resulting model files was predominantly correct, with minimal errors or ‘hallucinations’ observed, even when repeating the same prompt. Figure 2 illustrates an example where the AI modeling assistant was prompted to generate a model file based on a description of a Susceptible-Infected-Susceptible (SIS) system. The assistant not only produced an EMODL file with syntactically correct format but also accurately captured the mathematical specifics of the SIS model. It is important to note that the SIS model was not included in the initial few-shot learning used for prompt engineering. Interestingly, the AI assistant named the EMODL file simple-SIS and correctly described it as an SIS model in the response. This highlights that information about disease models and concepts likely existed in the corpus of documents used to train GPT4.

**Figure 2:**
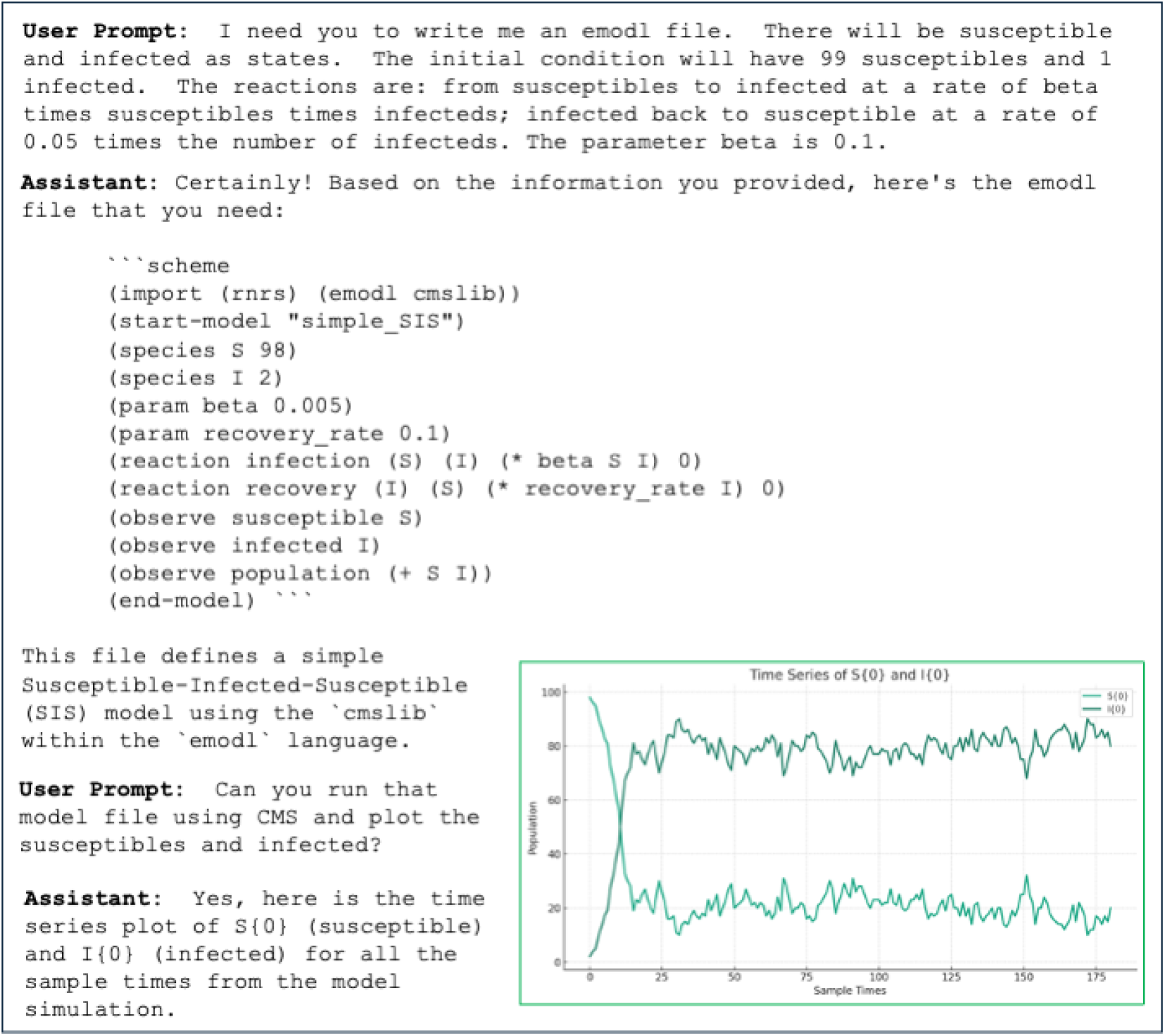
Interaction and output of the AI Modeling Assistant using the EMODL language for a Susceptible-Infected-Susceptible (SIS) disease model. The upper section of the image displays a text-based dialogue where the user requests the creation of an EMODL file, which the assistant generates based on specified model parameters and dynamics. Below, the assistant runs the model using a computational simulation engine (CMS) and presents a time series plot of the simulation results. This plot depicts the changes in the numbers of susceptible (S) and infected (I) individuals over time, demonstrating the model’s dynamics as simulated by the assistant. The dialogue showcases the assistant’s capabilities in model creation, execution, and result visualization, highlighting its practical application in disease modeling workflows.

### 3.3 The AI assistant can extract information from a model description in a document and translate to the EMODL format

Alternatively to direct input in a chat interface, users can upload a model description document to the AI assistant. This assistant is capable of extracting text from the document and converting the extracted model description into EMODL format. The nature of the model descriptions in these documents often mirrors those found in peer-reviewed journal articles on modeling; for an example of such a model description, refer to Appendix D.

This capability to directly convert textual model descriptions from documents into EMODL using natural language prompts represents a significant advancement in streamlining disease modeling workflows. However, there are notable limitations to this method. The assistant struggles with documents that are complex either in terms of their length, content, or if they are a scanned copy of an older document. A specific instance of this limitation is illustrated by the assistant’s inability to accurately extract the epidemiological model from the seminal 1996 research article by Eichner and Dietz (23). This document poses considerable challenges for parsing, as the model is described using a combination of text, tables, appendices, and equations, with the equations expressed in deterministic ordinary differential equation form even though the article describes simulating a discrete stochastic reaction network. While the AI assistant can extract portions of the information, it often requires additional input to accurately complete the model description.

### 3.4 The AI assistant can efficiently call CMS and plot the output from natural language prompts

The AI assistant is equipped to automatically initiate the execution of the CMS container using the model file alongside a standard configuration file. Upon completion of the simulation within the CMS container, the resulting data are uploaded into the AI assistant thread. Utilizing the Code Interpreter functionality, users can pose natural language questions to analyze and visually represent the data. Figure 2 provides an illustration of this capability, showcasing a request to plot the simulation outputs. In this example, the trajectories of susceptible and infected populations are graphically depicted over time, representing a minimal output from a disease model simulation.

This example, while illustrative, represents a simple model and scenario. More realistic modeling efforts may generate outputs that are significantly more complex and larger in size, potentially encompassing detailed statistical analyses such as calculating 95% confidence intervals for observable measurements across multiple model realizations. It is anticipated that there may be limitations to the volume of data that Code Interpreter can handle efficiently. To address larger datasets, the AI assistant has the capability to generate analytical scripts in languages such as python and R, facilitating offline data processing.

### 3.5 The AI assistant can navigate natural language conversations adapting a model iteratively

The AI assistant demonstrates the capability to iteratively refine model descriptions in collaboration with users. Figure 3 presents an example typical of infectious disease modeling, where the transmission characteristics differ between adults and children, such as higher transmission rates among children in group childcare settings. The assistant efficiently generated a new EMODL file that extended the model to include different age groups, both younger and older individuals. While this model was syntactically correct and compatible with the CMS simulation engine, the initial model description inaccurately omitted transmission routes between the designated age groups. This oversight could be considered either a ‘hallucination’ or the result of an inadequately specified prompt. Upon receiving a follow-up prompt to correct this omission (as shown in Figure 3), the assistant addressed the issue effectively.

**Figure 3:**
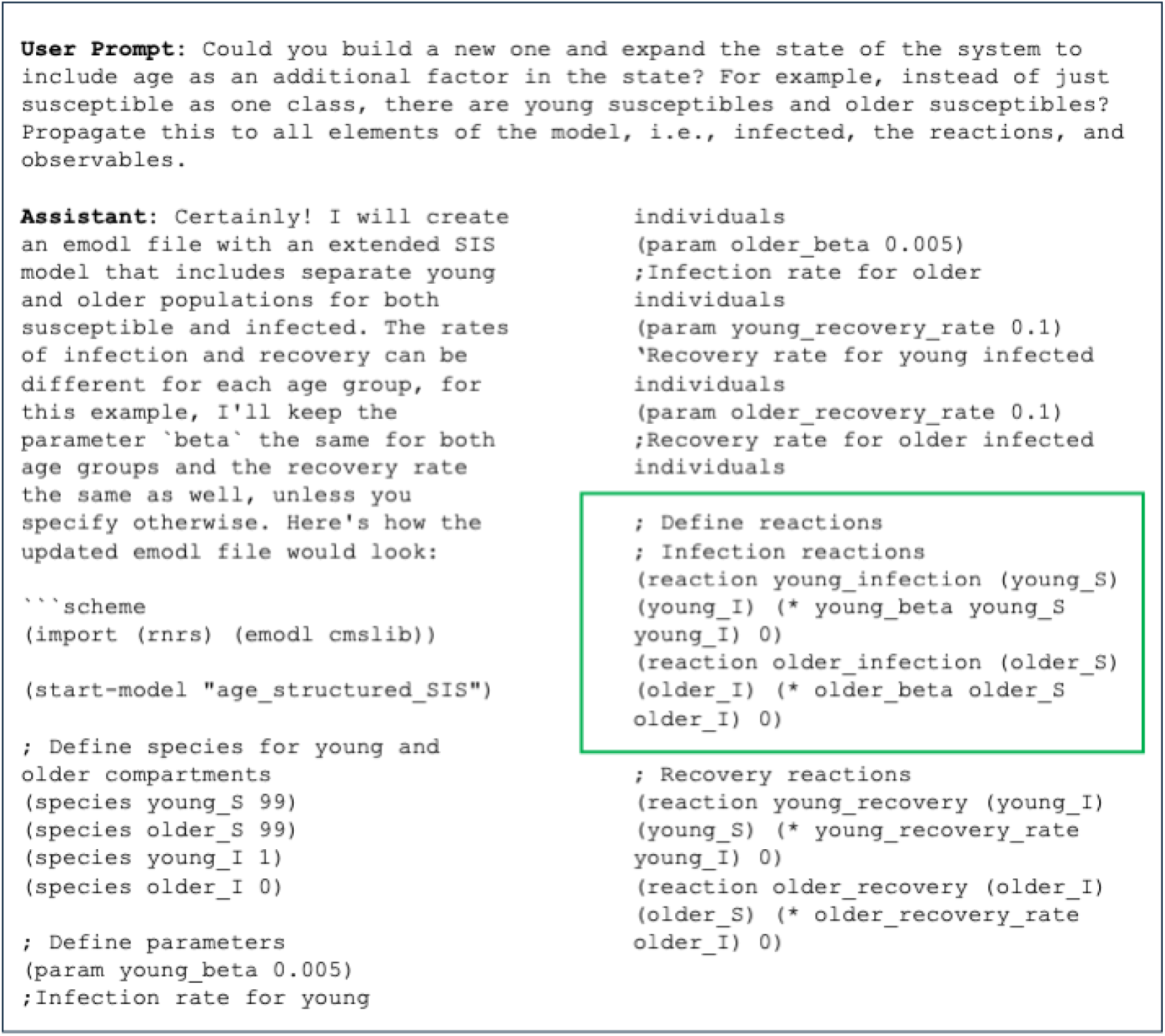
Enhancement of an EMODL file to include age stratification in a Susceptible-Infected-Susceptible (SIS) model. The figure depicts an exchange where the user requests the inclusion of age as a factor in the disease model, leading to separate categories for young and older individuals. The assistant generates a new model named “age structured SIS,” specifying separate compartments and infection dynamics for young and older populations. Parameters and reactions are defined for each group, with the initial conditions setting the young and older susceptible populations at 99, and infected populations at 1. Notably, the green box highlights a critical omission in the model: the lack of reactions representing transmission between the young and older groups.

For users with limited experience in model building, this highlights a critical limitation. To mitigate such issues, several strategies could be employed: expanding the range of example model files to cover various types of models, incorporating disease model building best practices into prompt engineering for a more interactive experience, and involving specialized agents to review the model files for common errors. These measures would enhance the reliability and user-friendliness of the AI assistant in complex model generation scenarios.

## 4 Discussion

This study aligns with contemporary efforts within the broader scientific and technological community, where the modular components of OpenAI’s offerings are increasingly being integrated into diverse workflows (24; 25; 26; 27). Particularly, functionalities such as file retrieval, Code Interpreter, and custom functions are pivotal in enhancing the capabilities of various applications (28). In application development, there is a growing trend towards constructing sophisticated apps that leverage these custom elements and plug-ins. Additionally, the ability to augment these applications with knowledge through retrieval-augmented generation (RAG) exemplifies the advancements in how AI can be tailored to meet specific research and commercial needs (18). Our work leverages these developments, demonstrating how AI assistants can be effectively applied in the domain of infectious disease modeling.

To our knowledge, the application of generative AI tools within the field of global health, as demonstrated in this study, represents a novel integration of cutting-edge AI technologies with traditional disease modeling. Prior initiatives have not fully harnessed the capabilities of large language models (LLMs) to enhance disease modeling processes. This work uniquely combines the strengths of AI, particularly its proficiency in text processing, summarization, and software development, with established and vetted numerical methodologies and software packages for disease modeling. The AI’s ability to generate syntactically correct model files from natural language inputs, contextualize the specific requirements of the disease being modeled, and iteratively refine these models through user interaction, highlights a significant advancement. Conversely, the mathematical modeling community has excelled in numerically integrating diverse models using sophisticated mathematical techniques and maintaining control over computational environments. By bridging these two domains, our approach circumvents the need for AI to redundantly develop numerical schemes that are already well-established. Instead, it leverages these existing strengths, providing a pathway to accelerate learning and adoption in disease modeling for researchers and policy makers in low and middle income countries.

## 5 Limitations and challenges

Despite the promising advancements demonstrated in this study, there are inherent limitations and challenges associated with the integration of AI in disease modeling. This study has highlighted the ability for AI to support disease modeling through building a working prototype, however, the application of the prototype has been on a small number of models for demonstration purposes; more testing is required to verify and validate the capabilities of the assistant and solicit feedback from the modeling community. In future iterations of this tool, we foresee a significant challenge is the AI’s capacity to accurately handle large, complex models. While simpler models have been successfully explored within this article, more intricate systems may not be as readily processed by the AI. To mitigate this issue, breaking down complex models into smaller, manageable components, employing multiple AI agents, and iteratively refining the outputs can enhance accuracy and feasibility.

Additionally, the Code Interpreter component, particularly in terms of data plotting, is not yet perfected. Challenges arise especially when handling large datasets, which may lead to suboptimal visualization outcomes. While improvements in these functionalities are anticipated as technology advances, providing supplementary code that can be executed directly in a browser may offer a more robust solution for handling complex analytical tasks. Furthermore, it is crucial to recognize that the efficacy of the workflow in generating accurate outputs can vary, and the responsibility for verifying and refining these outputs ultimately rests with the user. Acknowledging and addressing these challenges is essential for effectively leveraging AI tools in disease modeling and for ensuring that the results are both reliable and actionable.

## 6 Conclusion

Looking ahead, the prototype introduced in this study holds promising avenues for further evolution and application. One exciting direction is the expansion of the EMODL model file repository to include a greater variety of model types, each accompanied by text descriptions that elucidate their design and function. These models could be adaptively selected using RAG, tailored specifically to the requirements of the desired disease model, geographical context, and demographic considerations. Data to inform parameters and initial conditions could also be automatically retrieved through RAG or other retrieval methods. Moreover, the inclusion of multiple modeling and simulation engines, both bespoke and tailored, is feasible as long as these models are developed within a controlled computational environment such as Docker. The AI modeling assistant would then be able to select from a whole library of models allowing the tool to dynamically respond to diverse user needs and efficiently integrate new modeling platforms. The architecture required to achieve this broader vision of a modeling assistant will likely require multiple agents specialized to different sub-tasks with a coordinating agent. More generally, this conceptual architecture could be applied to a diverse set of modeling domains beyond infectious disease modeling and epidemiology.

The tool described in this article could be enhanced to incorporate a deeper knowledge of disease modeling by leveraging retrieval-augmented generation of scientific articles or best-in-class guidance on model construction, operation, and analysis. Such advancements would not only refine the tool’s utility but also broaden its appeal to a wider audience, particularly those lacking access to specialized tools and expertise. This could make the AI assistant an invaluable resource, especially in low- and middle-income countries, where it could significantly support public health decision-making by democratizing access to advanced disease modeling capabilities.

## Data Availability

There was not primary data produced by this study.

## A Assigning the AI modeling assistant’s role

The following is the assigned role for the modeling assistant:

You are an assistant helping to build model files to be executed by the compartmental modeling software CMS and plot the output of the model.

For the input, you will interact with the user to ask questions of the model or help parse a document that might describe the model. You will use Code Interpreter to output the model in the EMODL format.

Once you have written model, then you will call the CMS function to run the simulation engine on the model.

Then you will import the output file called trajectories.csv in order for the user to ask questions of the output and plot it in various ways.

## B Few Shot Learning

The following example EMODL files are included in the prompt for few shot learning. These are directly included as example files in the prompt for the AI Modeling assistant. These models can also be found in the source code repository ():

### B.1 Susceptible Exposed Infected Recovered (SEIR) Model

(import (rnrs) (EMODL cmslib))

(start-model “seir”)

(species S 990)

(species E 0)

(species I 10)

(species R 0)

(species CI 0)

(param beta 0.52)

(param Kei 0.25)

(param Kir 0.2)

(param Kwaning 0.0027397260273972603)

(reaction transmit (S) (E CI) (/ (* beta S I) (+ S E I R)) 0)

(reaction shed (E) (I) (* Kei E) 0)

(reaction recover (I) (R) (* Kir I) 0)

(reaction waning (R) (S) (* Kwaning R) 0)

(observe S S)

(observe E E)

(observe I I)

(observe R R)

(observe cumulative CI)

(observe population (+ S E I R))

(end-model)

### B.2 Garki model for malaria

(import (rnrs) (EMODL cmslib))

(start-model “garki.EMODL”)

(species X1 1000)

(species X2)

(species Y1 100)

(species Y2)

(species Y3)

(species X3)

(species X4)

(func totalpop (sum X1 X2 X3 X4 Y1 Y2 Y3))

(observe susceptible X1)

(observe latent X2)

(observe infected Y1)

(observe recovering Y2)

(observe immune (+ X3 X4 Y3))

(observe totalpopulation totalpop)

(param delta 0.0001) ; birth and death rate (param a 0.3) ; human biting rate

(param N 15) ; incubation in human

(param n 10) ; incubation in mosquito

(param alpha2 0.00019) ; rate of transitioning to fast recovery

(param g 0.097) ; susceptibility to bite

(param r1 0.0023) ; slow daily recovery rate

(param r2 0.023) ; fast daily recovery rate

(param alpha1 0.002) ; rate of losing infectivity

; seasonal parameter

(func C (* 0.2 (+ 1.01 (sin (* (/ time 365) 2 pi)))))

; infection rate

(func h (* g (- 1 (exp (/ (* (- C) Y1) totalpop)))))

(reaction birth () (X1) (* delta totalpop))

reaction deathX1 (X1) () (* X1 delta))

(reaction deathX2 (X2) () (* X2 delta))

(reaction deathX3 (X3) () (* X3 delta))

(reaction deathX4 (X4) () (* X4 delta))

(reaction deathY1 (Y1) () (* Y1 delta))

(reaction deathY2 (Y2) () (* Y2 delta))

(reaction deathY3 (Y3) () (* Y3 delta))

(reaction infectX1 (X1) (X2) (* X1 h))

(reaction latencyX2 (X2) (Y1) (* X2 100))

(reaction lossinfectY1 (Y1) (Y2) (* Y1 alpha1))

(reaction acquireimmunityY2 (Y2) (Y3) (* Y2 alpha2))

(reaction recoveryY3 (Y3) (X3) (/ (* Y3 h) (- (exp (/ h r2)) 1)))

(reaction infectX3 (X3) (X4) (* X3 h))

(reaction latencyX4 (X4) (Y3) (* X4 100))

(reaction recoveryY2 (Y2) (X1) (/ (* Y2 h) (- (exp (/ h r2)) 1))) (end-model)

### B.3 Polio model with surveillance

; polio_observation_model

(import (rnrs) (EMODL cmslib))

(start-model “polio_surveillance.EMODL”)

(species S 2500)

(species I 5)

(species Cu)

(species Cd)

(species R)

(species V)

(func population (sum S I Cu Cd R V))

(func cases (+ Cu Cd))

(func vaccinated (/ V population))

(observe vaccinated_ratio vaccinated)

(observe infection_ratio (/ I population))

(observe cases_ratio (/ cases population))

(observe susceptible_ratio (/ S population))

(observe recovered_ratio (/ R population))

(observe population population)

(observe susceptibles S)

(observe infections I)

(observe cases cases)

(observe observations Cd)

(observe recovereds R)

(observe vaccinated V)

(param mu1 0.005) ;birth rate

(param mu2 0.005) ;death rate

(param alpha (/ 1 200)) ;case/infection rate

(param gammaC (/ 1 30)) ;recovery rate cases

(param gammaI (/ 1 30)) ;recovery rate infections

(param betaC (/ 1 1000)) ;infectivity rate cases

(param betaI (/ 1 8000)) ;infectivity rate infections

;(param theta .01) ;RI vaccination rate less than 1

(param detection .8) ;active surveillance sensitivity

;parameters for vaccination outbreak response

(param x1 .25) ;upper asymptote is also the maximum coverage level for outbreak response

(param x2 0) ;lower asymptote

(param x3 1)

(param x4 1)

(param x5 .5) ;speed of response/growth rate

(param x7 3)

(param x8 0) ;vertical shift

;infection-triggered

;x6 between 80-120 produces multiple vaccination events when paired with alpha 1 over 200

;274 approximates 250 threshhold

;524 approximates 500 threshhold

;(param x6 600) ;median horizontal shift point which is somewhat higher than the trigger point

;case detection-triggered

;use vaccination campaign calculation spreadsheet to estimate parameter x6

;44 approximates 20 threshhold

;39 approximates 15 threshhold

;34 approximates 10 threshhold

;29 approximates 5 threshhold

(param x6 34) ;median horizontal shift point which is somewhat higher than the trigger point

(reaction birth () (S) (* mu1 population))

(reaction deathS (S) () (* mu2 S))

(reaction deathV (V) () (* mu2 V))

(reaction deathI (I) () (* mu2 I))

(reaction deathCu (Cu) () (* mu2 Cu))

(reaction deathCd (Cd) () (* mu2 Cd))

(reaction deathR (R) () (* mu2 R))

(reaction infectI (S) (I) (* betaI S I))

(reaction infectC (S) (I) (* betaC Cu))

(reaction cases (I) (Cu) (* alpha I))

(reaction recoveryI (I) (R) (* gammaI I))

(reaction recoveryCu (Cu) (R) (* gammaC Cu))

(reaction recoveryCd (Cd) (R) (* gammaC Cd))

(end-model)

## C Example model file from zero-shot learning assistant

The following is the exact user prompt and model output from the assistant in zero shot learning:

**User**: I need you to write me an EMODL file. There will be susceptible and infected as states. The initial condition will have 99 susceptibles and 1 infected. The reactions are: from susceptibles to infected at a rate of beta times susceptibles times infecteds; infected back to susceptible at a rate of 0.1 times the number of infected individuals. The parameter beta is 0.005.

**Model produced by assistant in code:**

;; Define compartments

(compartments

(susceptible)

(infected)

)

;; Set initial conditions

(initial

(susceptible 99)

(infected 1)

)

;; Define parameters

(parameters

(beta 0.005)

(gamma 0.1)

)

;; Define reactions

(reactions

;; Infection process

(reaction infection

(reactants (susceptible 1) (infected 1))

(products (infected 2))

(rate (* beta susceptible infected))

)

;; Recovery process

(reaction recovery

(reactants (infected 1))

(products (susceptible 1))

(rate (* gamma infected))

)

)

## D Model description in a separate document

In this section, we provide the model description that is contained in a pdf that is uploaded to the AI Modeling Assistant using the file retrieval functionality. This file can also be found in the code repository ().

### D.1 Model Description

#### D.1.1 States of the model and initial condition for simulation

The following is a list of the states of the model:

- Susceptibles: 100
- Infected: 1

#### D.1.2 Parameters of the model

The following is a list of the parameters and their values in the model:

- *β* = 1
- *γ* = 0.1

#### D.1.3 Reactions of the model

The following is a list of the stochastic reactions in the model:

- Susceptibles → Infected, at a rate of *β* × Susceptibles × 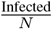
- Infected → Susceptibles, at a rate of *γ* × Infected

#### D.1.4 Observables of the model

The following are the observables of the system:

- Susceptibles
- Infected
- Total Population which is Susceptibles + Infected

